# Predicting Depressive Symptoms Among Reproductive-Aged Women in Bangladesh Using Bagging Ensemble Machine Learning on Imbalanced Bangladesh Demographic and Health Survey 2022 Data

**DOI:** 10.64898/2026.04.22.26351445

**Authors:** Sultan Mahmud, Mt Sonia Akter, Bibek Ahamed, Ahmed Ehsanur Rahman, Shams El Arifeen, Aniqa Tasnim Hossain

## Abstract

**Background:** Depressive symptoms among reproductive-aged women represent a major public health concern in low- and middle-income countries, yet systematic screening remains limited. In most population survey datasets, the low prevalence of depression results in severe class imbalance, which challenges conventional machine learning models. Therefore, we develop and evaluate a bagging-based ensemble machine learning framework to predict depressive symptoms among reproductive-aged women using highly imbalanced Bangladesh demographic and health survey (BDHS) 2022 data.

**Methods:** The sample comprised women aged 15–49 years drawn from BDHS 2022 data. Depressive symptoms were defined using the Patient Health Questionnaire (PHQ-9 ≥10). Candidate predictors were drawn from sociodemographic, reproductive, nutritional, psychosocial, healthcare access, and environmental domains. Feature selection was performed using Elastic Net (EN), Random Forest (RF), and XGBoost model. Five classifiers (EN, RF, Support Vector Machine (SVM), K-nearest neighbors (KNN), and Gradient Boosting Machine (GBM)) were trained using both oversampling-based approaches and the proposed ensemble framework. Model performance was evaluated on an independent test set using accuracy, sensitivity, specificity, F1-score, and the normalized Matthews correlation coefficient (normMCC).

**Results:** Approximately 4.8% of women were identified with depressive symptoms. The proposed bagging ensemble framework consistently achieved more balanced predictive performance than oversampling-based models. Average normMCC improved from 0.540 (oversampling) to 0.557 (ensemble). RF and GBM ensembles demonstrated notable improvements in identifying depressive cases, while the EN ensemble achieved the highest overall performance and sensitivity. Threshold optimization yielded stable normMCC across models, indicating robust trade-offs between sensitivity and specificity.

**Conclusions:** Bagging-based ensemble learning provides a more robust and balanced approach than synthetic oversampling for predicting depressive symptoms in highly imbalanced population survey data. This approach has important implications for improving early identification and population-level mental health surveillance in resource-constrained settings.

## Introduction

Depression is a leading contributor to the global burden of disease and one of the primary causes of disability worldwide [1], with women of reproductive age bearing a disproportionate share of this burden [2]. In low- and middle-income countries (LMICs), depressive symptoms among women are closely linked to socioeconomic disadvantage, reproductive and maternal health challenges, gender inequality, and limited access to mental health services [2, 3]. Despite increasing recognition of women’s mental health as a public health priority, depression remains substantially underdiagnosed and undertreated in many LMIC settings, including Bangladesh [4–6].

In Bangladesh, existing evidence indicates a considerable prevalence of depressive symptoms among women; however, systematic screening and early identification remain limited [5]. This gap reflects shortages of trained mental health professionals, persistent stigma, and weak integration of mental health services into primary healthcare systems [4]. In this context, large-scale population-based surveys such as the Bangladesh Demographic and Health Survey (BDHS) offer a valuable opportunity to identify women at elevated risk of depression using routinely collected sociodemographic, reproductive, and household-level indicators [7]. Leveraging these nationally representative datasets for predictive modeling could enable scalable, cost-effective strategies for population-level mental health surveillance in resource-constrained settings.

Machine learning (ML) methods have been increasingly applied in mental health research to improve risk prediction and capture complex, nonlinear relationships among diverse predictors [8–10]. Population-based surveys, including Demographic and Health Surveys (DHS), have also been used to identify high-risk subgroups and support mental health surveillance in LMICs [11–13]. However, applying ML to DHS-type data presents distinct methodological challenges. First, depressive symptoms are relatively rare in general population surveys, resulting in extreme class imbalance [7], where the minority (depressed) class constitutes a small fraction of the sample. Second, DHS data are characterized by heterogeneous socio-demographic and environmental variables with limited clinical information, which further complicates predictive modeling. In such settings, conventional ML models often achieve high overall accuracy while performing poorly in identifying true cases, thereby limiting their usefulness for screening and early intervention.

Several approaches have been proposed to address class imbalance, including under-sampling and over-sampling strategies [14, 15]. While these methods attempt to rebalance class distributions, synthetic over-sampling techniques such as the Synthetic Minority Oversampling Technique (SMOTE) may introduce artificial patterns, distort the underlying epidemiological structure, and reduce generalizability—particularly in population-based survey data where preserving the original data distribution is critical [16]. These limitations highlight the need for alternative approaches that can improve minority-class detection without compromising data integrity.

Ensemble learning methods, particularly bagging (bootstrap aggregating), offer a promising solution for modeling rare outcomes in epidemiological data. By training multiple models on different subsets of the data—where class distributions can be balanced through resampling—bagging increases exposure to minority cases while reducing variance and overfitting [16, 17]. Importantly, unlike synthetic over-sampling, bagging preserves the original data structure, making it well suited for DHS-type datasets. Furthermore, combining ensemble learning with threshold optimization allows flexible control over the trade-off between sensitivity and specificity, which is essential in public health applications where the cost of false negatives and false positives may differ [18].

In addition to modeling strategies, the choice of evaluation metric is critical in imbalanced classification. Common metrics such as accuracy and AUC can provide misleadingly optimistic assessments in highly imbalanced settings [16]. The Matthews correlation coefficient (MCC), particularly in its normalized form (normMCC), offers a more reliable and balanced measure by incorporating all elements of the confusion matrix and providing a robust assessment of model performance under class imbalance[16].

In this study, we develop and evaluate a bagging-based ensemble machine learning framework tailored for highly imbalanced DHS data to predict depressive symptoms among reproductive-aged women in Bangladesh using BDHS 2022 data. We systematically compare the proposed ensemble approach with conventional oversampling-based models and assess performance using metrics appropriate for imbalanced classification, with a focus on normMCC. By integrating balanced-subset bagging with threshold-optimized decision rules, this study aims to advance methodological approaches for imbalanced epidemiological prediction while maintaining the integrity of survey data.

Beyond methodological contributions, this study has important policy implications. The proposed framework provides a scalable and data-driven approach for identifying high-risk women using routinely collected national survey data. At the community and primary care levels, it can support frontline health workers in prioritizing individuals for early screening and referral. At the program and national levels, it can inform resource allocation, targeted interventions, and integration into digital health and surveillance systems, thereby strengthening mental health service delivery in Bangladesh and similar resource-constrained settings.

## Methods

This study employed a structured machine learning workflow to predict depressive symptoms among reproductive-aged women using imbalanced Bangladesh Demographic and Health Survey (BDHS) 2022 data. The analytical pipeline comprised data preprocessing, supervised feature selection, model development, and performance evaluation (Fig. 1).

**Fig. 1.**
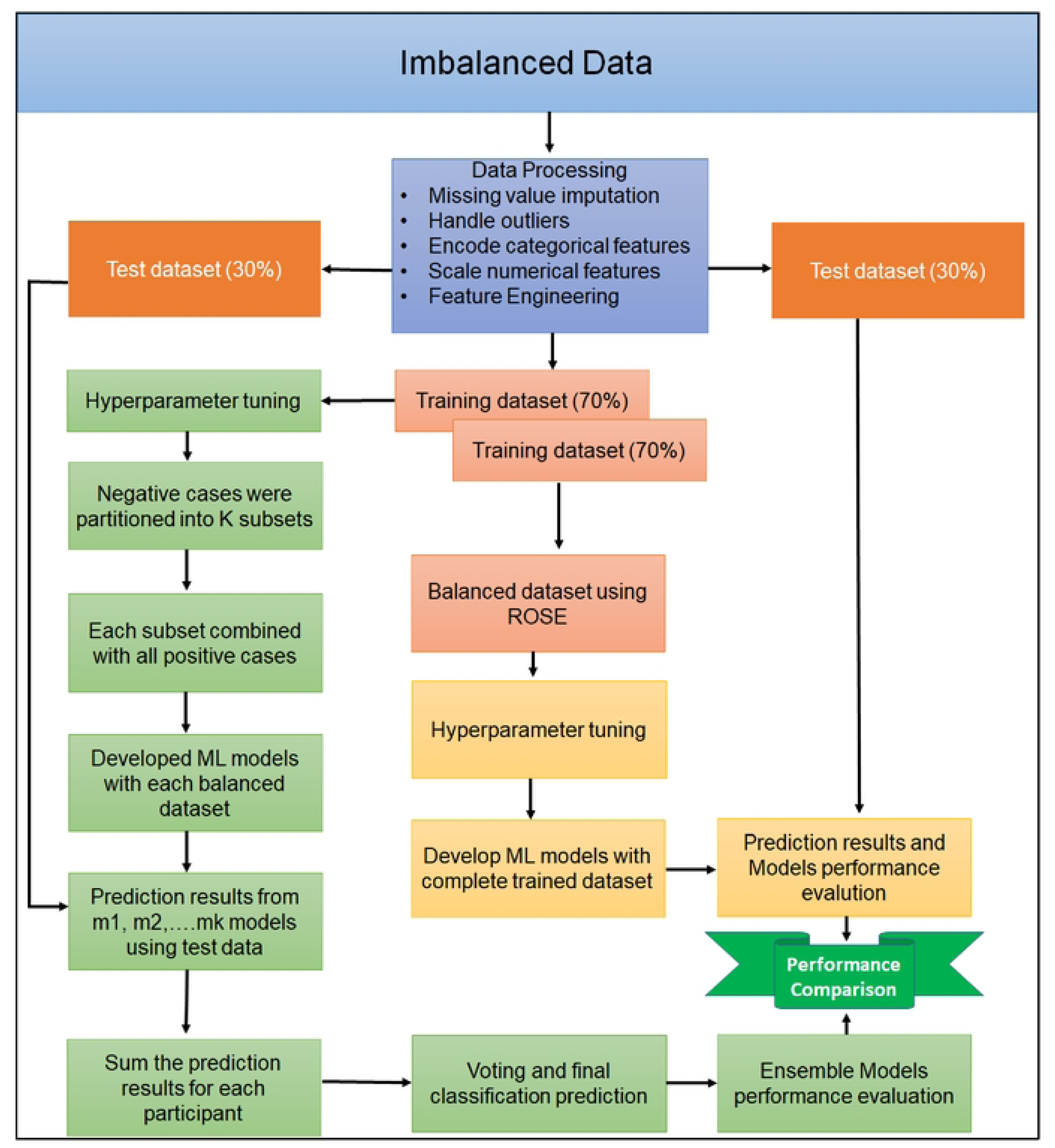
Workflow for predicting depressive symptoms among reproductive-aged women in Bangladesh using imbalanced BDHS data. The pipeline includes data preprocessing, Hyperparameter tuning, training multiple classical machine learning models, constructing bagging ensembles, and evaluating model performance on the test set.

### Study Design and Data Source

This study employed a cross-sectional design using data from the BDHS 2022, a nationally representative survey that collects information on health, nutrition, and socio-demographic characteristics of women aged 15–49 years. The BDHS uses a two-stage stratified cluster sampling design to ensure national representativeness.

The analysis included reproductive-aged women (15–49 years) with available data on depressive symptoms measured using the Patient Health Questionnaire (PHQ-9). Women with missing outcome data were excluded. The final analytic sample comprised 17,266 women.

### Outcome Variables

Depressive symptoms were assessed using the 9-item Patient Health Questionnaire (PHQ-9), a widely validated screening tool in Bangladeshi populations [7, 19, 20]. A binary outcome variable was defined using a cut-off score of ≥10 to indicate clinically significant depressive symptoms, while scores <10 were classified as non-depressed [7]. As a screening instrument, PHQ-9 reflects symptom burden rather than clinical diagnosis.

### Candidate Predictors

All candidate predictors were selected a priori based on existing literature on maternal mental health and data availability in the BDHS. Variables were grouped into the following domains (Fig. 2). Detailed coding and categorization of all predictors are provided in supplementary Table S1.

- Sociodemographic characteristics: age, education, residence, religion, marital status, geographic division, household wealth index (five quintiles), husband’s education, and household head characteristics
- Empowerment indicators: employment status, participation in household decision-making, and ownership of assets
- Maternal and reproductive factors: pregnancy status, pregnancy termination history, parity, presence of under-five children, and recent child death
- Nutritional and household characteristics: dietary diversity, sanitation, cooking facility, mobile financial use, livestock, and agricultural land ownership
- Psychosocial proxies: attitudes toward wife-beating and household crowding
- Healthcare access barriers: cost, distance, permission, transportation, and related barriers
- Environmental conditions: drinking water access and housing quality.

**Fig. 2.**
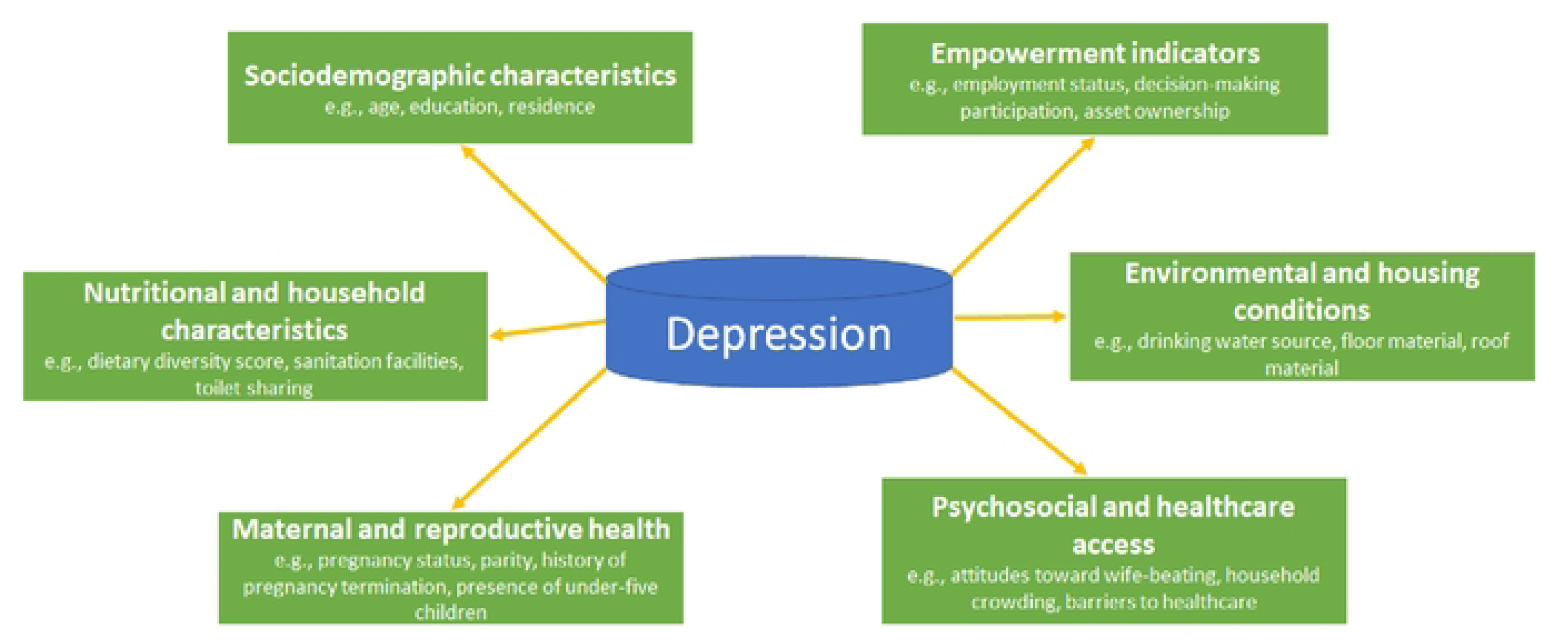
The features related to Depressive symptoms identified from BDHS data were grouped into categories

### Data Preprocessing

Data preprocessing was conducted prior to model development. Missing values were identified in two categorical variables: husband’s education (4.0%) and toilet sharing (0.27%). These were imputed using the K-nearest neighbors (KNN) method (k = 5) [21], which preserves multivariate relationships without assuming parametric distributions.

Categorical variables were encoded into appropriate numerical formats, and continuous variables were normalized using Min-Max scaling. These steps ensured that all features were standardized and suitable for machine learning algorithms.

### Splitting Data

The dataset was randomly partitioned into a training set (70%; n = 12,087) and a test set (30%; n = 5,179) [16], with the split stratified by outcome to preserve class imbalance. Two separate modeling pipelines were constructed using the same train–test split.

In the first pipeline, classical machine learning models were trained on the training data after addressing class imbalance using the Synthetic Minority Oversampling Technique (SMOTE). In the second pipeline, the bagging ensemble approach was developed using the original (non-oversampled) training data. For both pipelines, model performance was evaluated on the same independent test set, where no oversampling was applied.

### Feature Engineering

Feature selection was performed to simplify models, improve interpretability, reduce computational burden, and enhance predictive performance by retaining the most informative variables while minimizing overfitting [22]. Among different approaches, supervised feature selection methods generally demonstrate superior performance in predictive modeling [23]. Accordingly, this study employed a multi-model supervised feature selection framework using three widely applied techniques: Elastic Net (EN) [24], Random Forest (RF)[25], and Gradient Boosting Machine (GBM)[26]. The final model included predictors, defined as the union of features selected by the these feature selection techniques [27]. Feature selection was performed exclusively on the training dataset to avoid information leakage.

EN regularized logistic regression was applied to the full set of candidate predictors, combining L1 and L2 penalties to enable variable selection while accounting for multicollinearity. The mixing parameter was set to α = 0.5, and the optimal regularization parameter (λ) was selected using 10-fold cross-validation based on minimum deviance.

A RF model with 500 trees was used to estimate variable importance based on the mean decrease in Gini impurity. Similarly, an XGBoost model with a logistic objective function was trained, and feature importance was assessed using the gain metric.

To enhance robustness and reduce model-specific bias, the final feature set was defined as the union of predictors selected across the three methods. This approach ensured that variables consistently identified as important across different modeling frameworks were retained for subsequent analysis.

### Classical Machine Learning Model

To predict depressive symptoms, we evaluated a diverse set of machine learning models that capture different assumptions about data structure, nonlinearity, and feature interactions. The selected models represent complementary methodological families and are commonly used in health prediction research.

Elastic Net logistic regression (EN) [28] was included as a baseline interpretable model. By combining L1 and L2 regularization, EN effectively handles multicollinearity, performs embedded feature selection, and reduces overfitting in high-dimensional settings. Its coefficients provide direct interpretability, which is particularly valuable in public health applications.

Random Forest (RF) [29] was considered to address class imbalance in depressive symptoms outcomes. This ensemble tree-based approach captures complex nonlinear relationships and higher-order interactions while reducing bias toward the majority class through balanced resampling. RF models are robust to outliers and insensitive to feature scaling.

Support Vector Machine (SVM) [30] with a radial basis function kernel was included for its strong performance in nonlinear classification problems. The radial kernel allows SVM to model complex decision boundaries in high-dimensional feature spaces, making it suitable for heterogeneous survey data.

K-nearest neighbors (KNN) [31] was evaluated as a distance-based, nonparametric model that makes minimal assumptions about data distribution and serves as a useful benchmark for assessing local similarity structures.

Gradient Boosting Machine (GBM) [32] was included as a powerful ensemble method that sequentially combines weak learners to optimize predictive accuracy. GBM is well-suited for tabular health data, can model nonlinearities and interactions, and often achieves superior classification performance.

Models not included were Naïve Bayes (conditional independence assumption unlikely to hold for correlated socioeconomic and health variables), deep learning models (moderate sample size and limited interpretability), standalone decision trees (unstable and inferior to ensemble trees), and unregularized logistic regression (more prone to multicollinearity and overfitting than Elastic Net). Additionally, the XGBoost model was used for feature selection but excluded from final modeling due to instability during hyperparameter tuning, including convergence issues and highly variable performance across cross-validation folds, which limited its reliability and reproducibility in this setting.

### Bagging-Based Ensemble Learning Framework

For each classical model, a bagging ensemble was implemented using the training dataset, and performance was evaluated on the test set. Fig. 3 demonstrates the detailed process of the bagging ensemble. In this approach, K submodels were trained based on K subsets of the training data, where K was defined as the ratio of non-depressed to depressed cases in the training set (non-depressed/depressed). In this study, K=20.

**Fig. 3.**
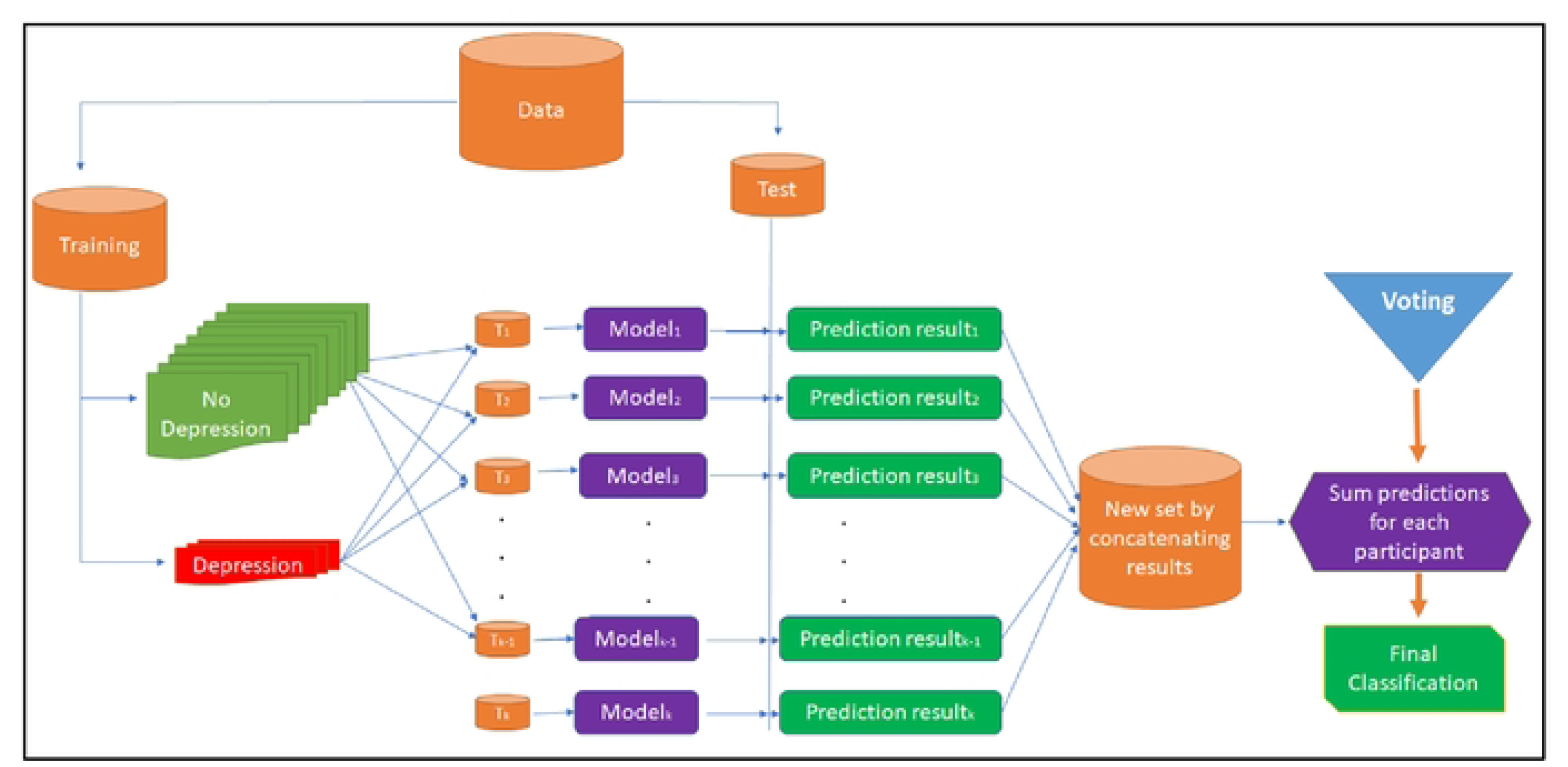
Pipeline for prediction of depressive symptoms among reproductive-aged women in Bangladesh using imbalanced BDHS data and an ensemble machine learning approach

To address class imbalance, records without depressive symptoms were partitioned into K non-overlapping subsets of approximately equal size (n ≈ 575), each comparable to the number of depressed cases. Each subset was then combined with all depressed cases to form balanced training sets (T_1_, T_2_,…,T_K-1,_ T_K_) containing 1,156 records in the first K−1 subsets and 1,157 records in the K^th^ subset. These balanced datasets were used to train K parallel submodels (Model_1_, Model _2,_…, Model_K-1,_ Model_K_) using the same base learner (Fig. 3).

During testing, each submodel generated predictions for all observations in the held-out test set. The predictions were aggregated by counting the number of submodels that classified an individual as depressed. A threshold-based voting rule was then applied to determine the final classification: an individual was labeled as depressed if the proportion of positive predictions exceeded a predefined threshold, ensuring consensus across the majority of submodels.

### Cross Validation and Hyperparameter Tuning

K-fold cross-validation (CV) is a commonly applied method in machine learning that offers a more reliable evaluation of model performance compared to a single train–test split [33]. In this approach, the dataset is divided into K equally sized folds. The model is trained and evaluated K times, with each fold serving once as the validation set while the remaining K−1 folds are used for training. The overall performance is then obtained by averaging the results across all folds. In this study, a value of K = 10 was used.

Hyperparameter tuning for all machine learning models was conducted using systematic search strategies in conjunction with cross-validation to identify the optimal parameter settings. A grid search approach was applied, whereby predefined ranges of candidate values were specified for each hyperparameter. All possible combinations within these ranges were explored, and model performance was evaluated for each configuration. The optimal values of the parameters for all models are presented in Table S2 (Supplementary file 2) and were consistently applied across both oversampling-based and ensemble modeling pipelines.

### Evaluation Metrics

Although accuracy, F1 score, and AUC are commonly used performance metrics for binary classification, their application to imbalanced datasets may result in overly optimistic evaluations. In contrast, the Matthews correlation coefficient (MCC) provides a more reliable and balanced assessment of predictive performance under class imbalance [16, 34]. Accordingly, model performance in this study was evaluated using accuracy, sensitivity, specificity, F1 score, and MCC (and its normalized form, normMCC), as defined in Equations (1)–(6).

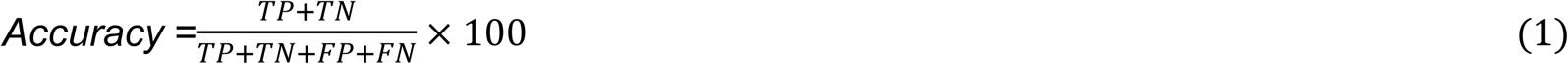

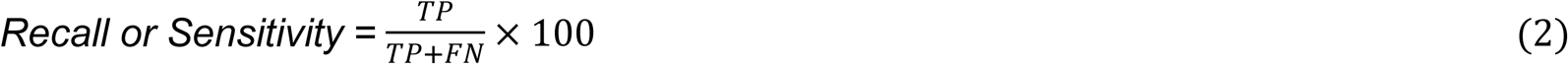

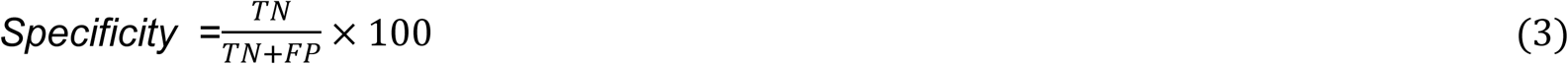

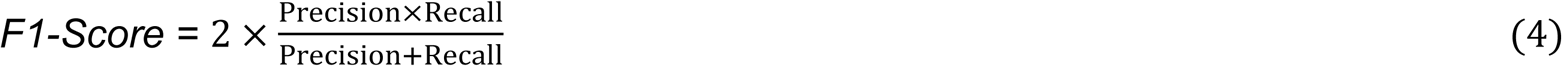

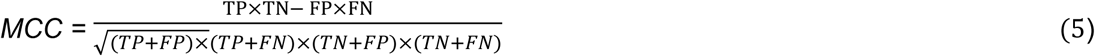

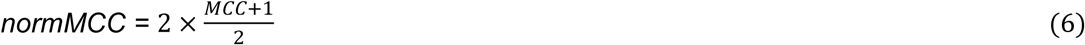

Where TP, TN, FP, and FN represent the numbers of true positives, true negatives, false positives, and false negatives, respectively. Precision is defined as the proportion of correctly predicted positive cases among all predicted positives (TP / (TP + FP)).

### Statistical Analysis

Descriptive statistics were used to summarize the characteristics of the study population. Continuous variables were reported as means and standard deviations or medians and interquartile ranges, as appropriate, while categorical variables were summarized using frequencies and percentages. Pearson’s chi-square (χ²) test and Student’s t-test were used to assess associations and differences between each predictor and depressive symptom status. Data management and statistical analyses were conducted using Stata and R (version 4.5.3), with model development implemented using the caret, glmnet, ranger, gbm, and xgboost packages.

Although the BDHS employs a complex survey design incorporating sampling weights, stratification, and clustering, these design features were not included in model training. This decision was made because the primary objective of this study was individual-level risk prediction rather than population-level inference. Machine learning algorithms are primarily designed to optimize predictive performance by learning patterns in the observed data, and the integration of survey weights into such models remains methodologically challenging and is not yet standardized across different algorithms and implementations.

Furthermore, prior methodological research suggests that the omission of sampling weights in predictive modeling often has limited impact on classification performance, particularly when models are evaluated using independent test data [35, 36]. In this context, unweighted models can still yield valid and generalizable predictions, as performance is assessed based on out-of-sample predictive accuracy rather than unbiased estimation of population parameters [36]. Therefore, consistent with existing literature on predictive modeling [12, 16, 33, 37], we prioritized model performance, stability, and generalizability over design-based inference.

### Ethical Considerations

The Bangladesh Demographic and Health Survey (BDHS) data are publicly available and fully de-identified. The original survey was conducted in accordance with national and international ethical standards, including procedures for informed consent, participant privacy, and confidentiality. This study is a secondary analysis of anonymized data and therefore did not require additional ethical approval. The data was accessed on Feb 10, 2026, and the authors did not have access to any information that could identify individual participants at any stage of the research. All analyses were conducted in compliance with established ethical guidelines for the use of publicly available data. The study was conducted in accordance with the TRIPOD-AI reporting guidelines [38] (Supplementary file 3).

## Findings

### Descriptive Statistics

A total of 17,266 women were included in the analysis, of whom 830 (4.81%) exhibited depressive symptoms. The mean age of respondents was 30.81 years (SD: 8.56), and the average years of education was 3.56 (SD: 1.54). Most participants resided in rural areas (63.87%) and identified as Muslim (89.84%). The majority were currently married (95.88%) and lived in male-headed households (85.51%) (Table 1).

**Table 1.**
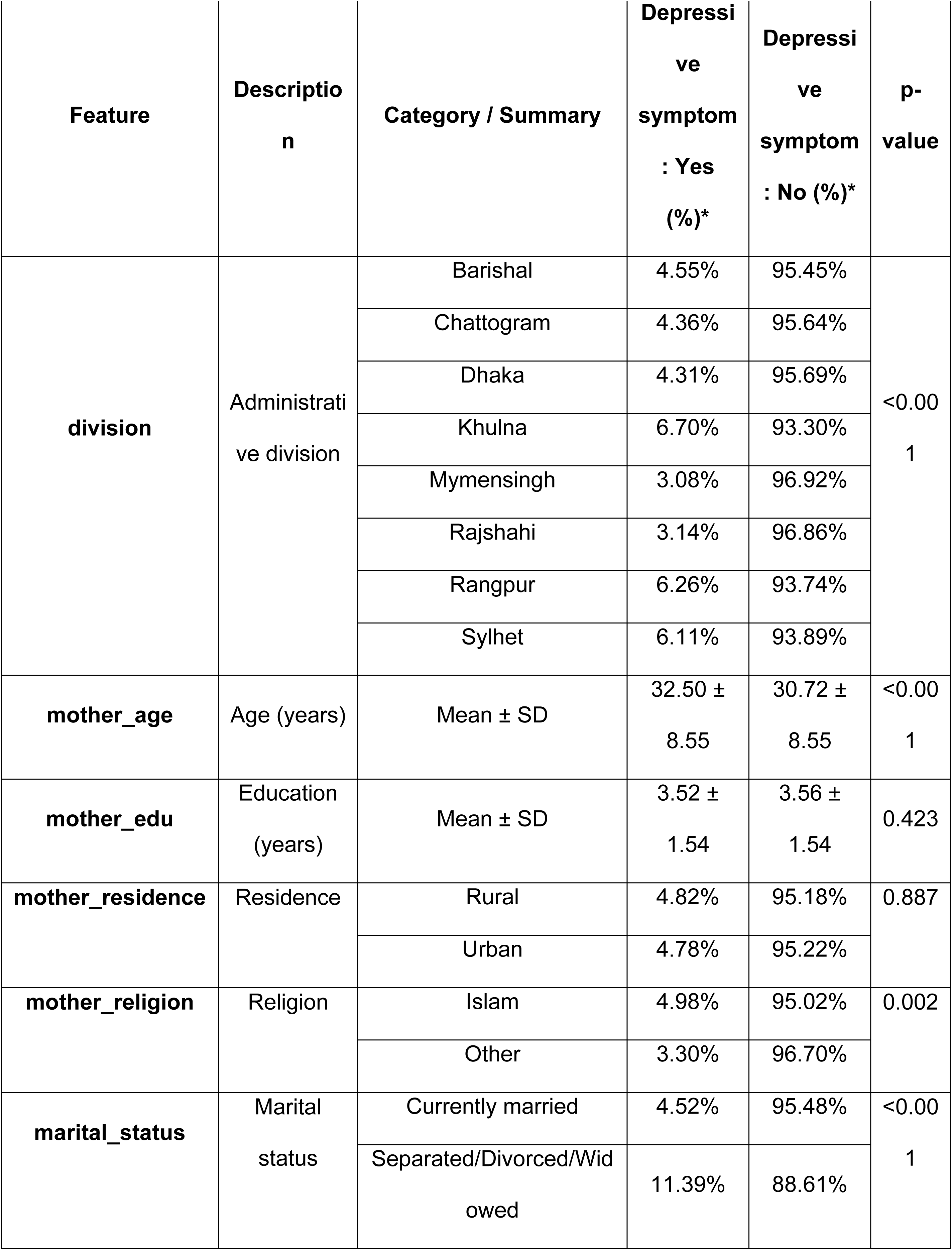
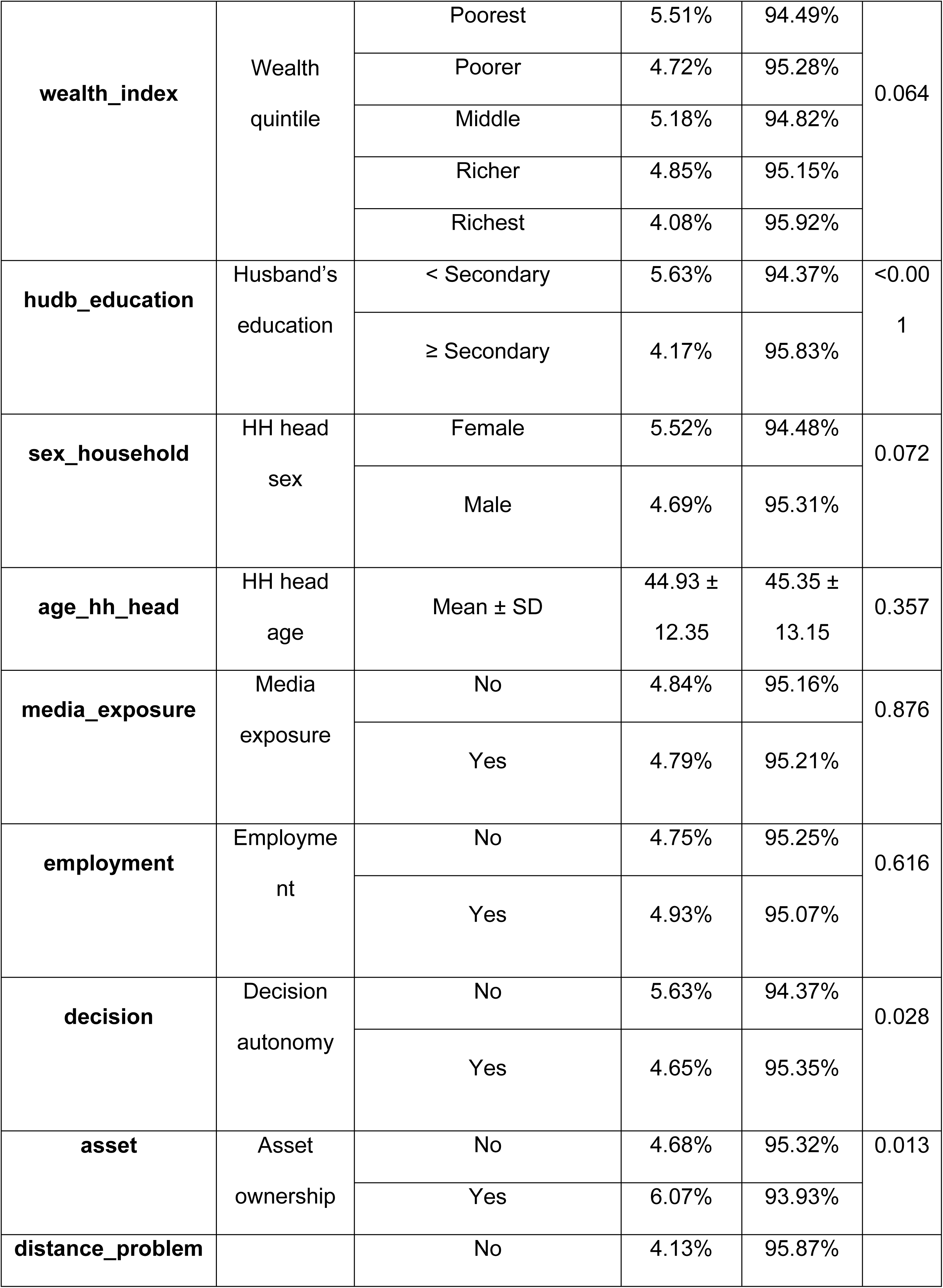
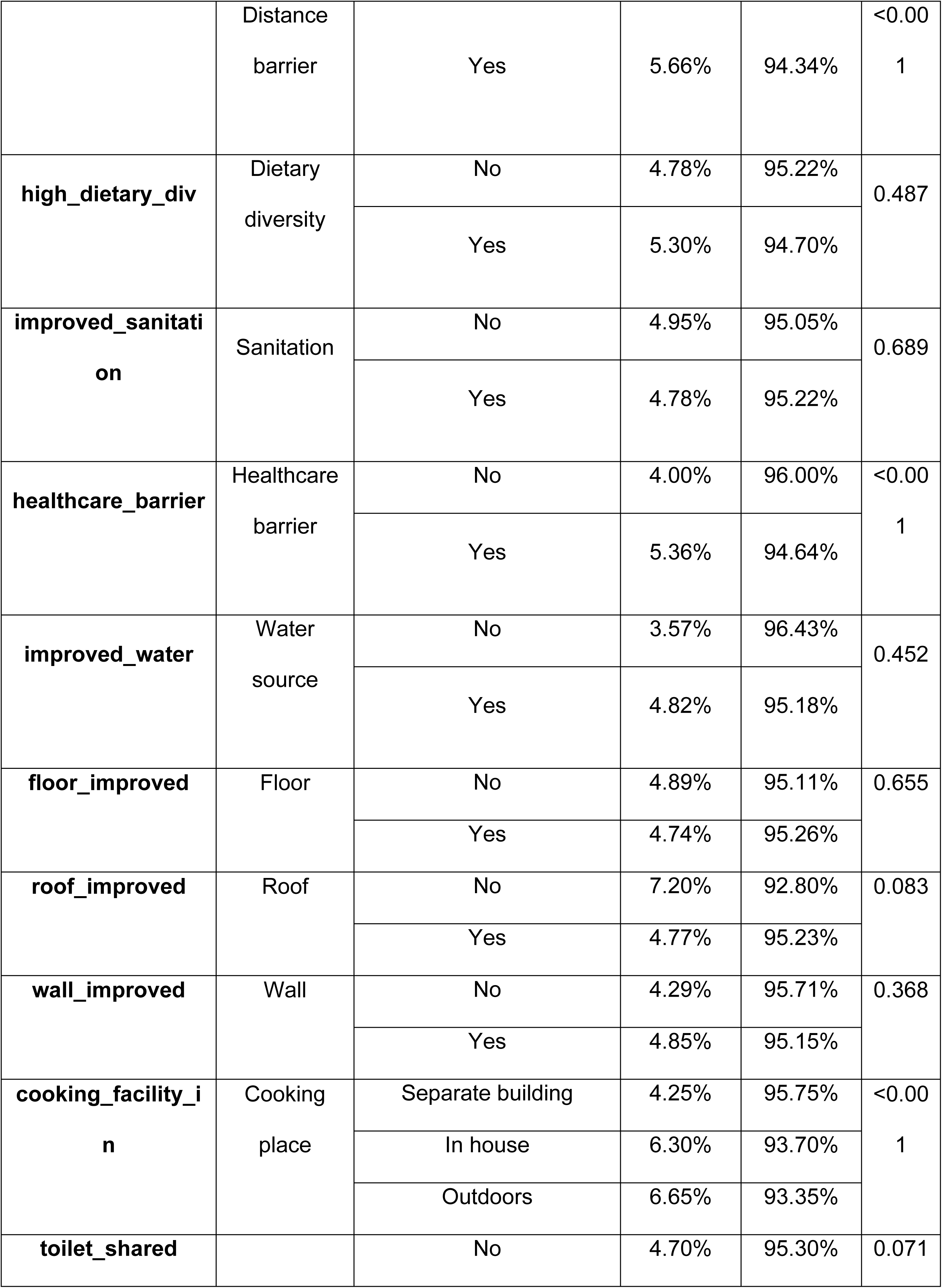
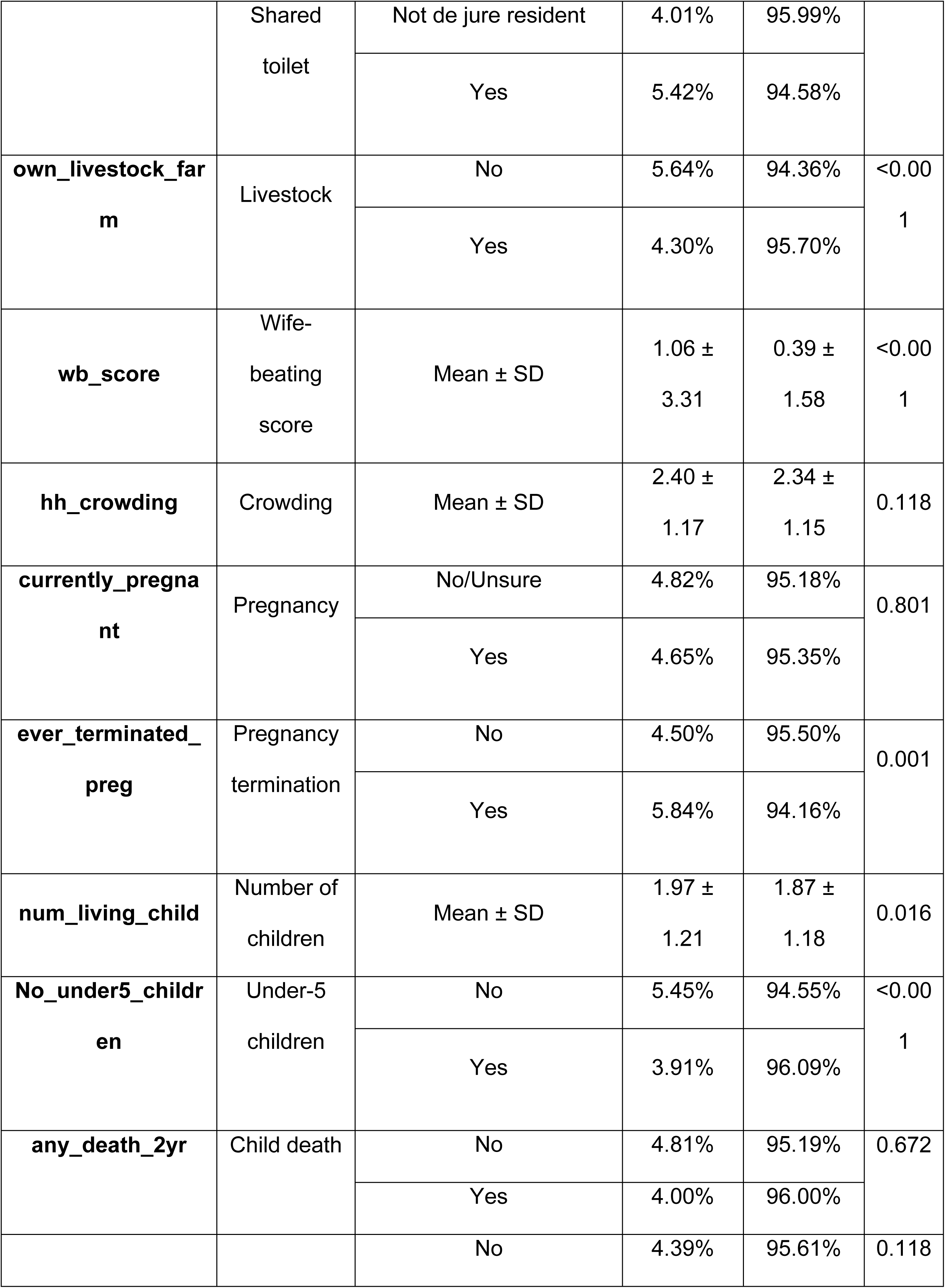
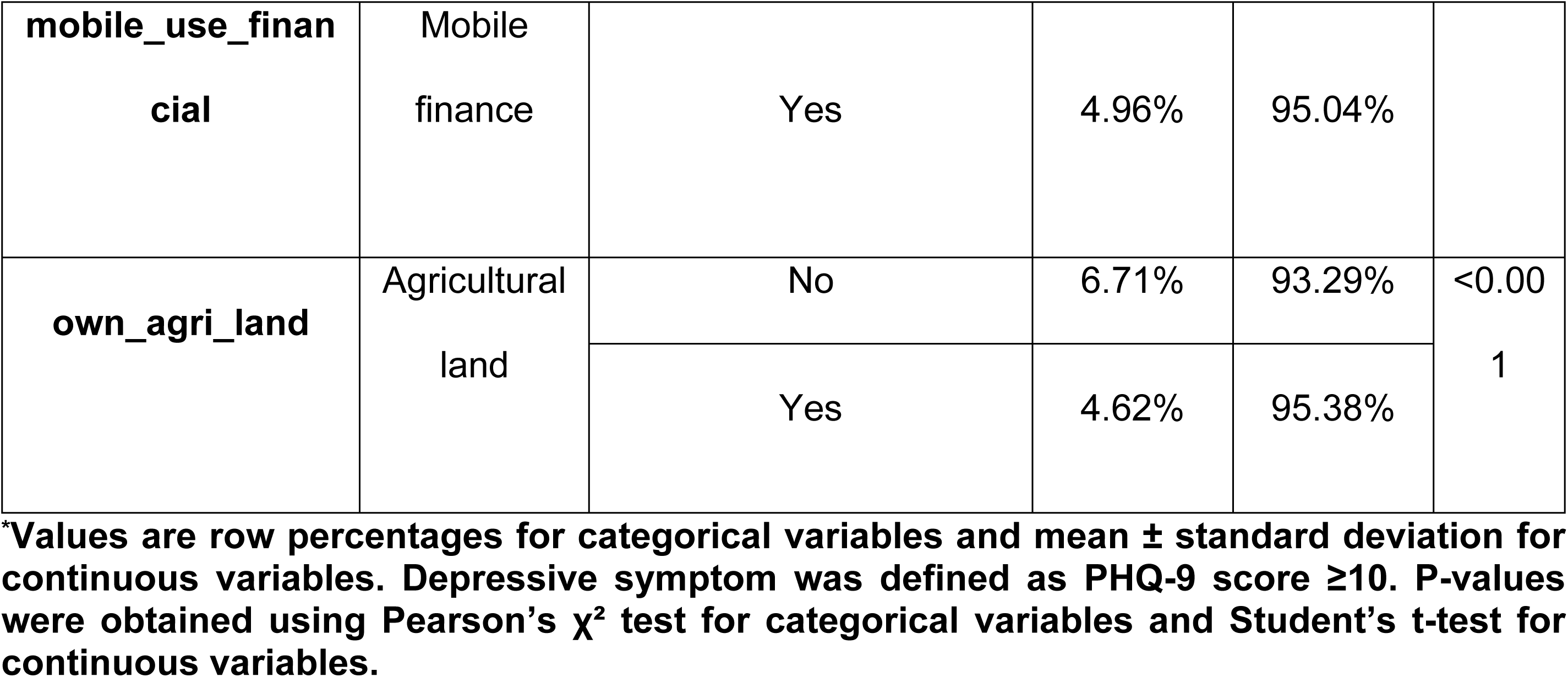
Socio-demographic, household, and reproductive characteristics of respondents by depressive symptom status (PHQ-9 ≥10) (N=17,266).

Socioeconomic characteristics showed that respondents were relatively evenly distributed across wealth quintiles, with the largest proportion in the richest group (24.02%). Approximately 56.59% of husbands had secondary or higher education. Around 61.46% of women were exposed to mass media, and 29.35% were employed. Most women reported having decision-making autonomy (83.95%), while only 9.26% owned household assets.

Regarding household and environmental conditions, 82.09% had access to improved sanitation and 99.03% had access to improved water sources. More than half (59.35%) reported barriers to accessing healthcare. The majority used a separate building for cooking (75.55%), and 62.43% owned livestock. The mean household crowding index was 2.34 persons per room (SD: 1.15).

In terms of reproductive characteristics, 6.48% of women were currently pregnant, and 23.02% had ever experienced pregnancy termination. The average number of living children was 1.88 (SD: 1.19), and 41.90% had at least one child under five years of age. Child mortality in the past two years was low (0.72%). Additionally, 72.83% of respondents reported using mobile financial services, and 90.94% owned agricultural land.

### Factors Associated with Depressive Symptoms

The prevalence of depressive symptoms varied significantly across several characteristics. Significant regional variation was observed (p < 0.001), with higher prevalence in Khulna (6.70%), Rangpur (6.26%), and Sylhet (6.11%), and lower prevalence in Mymensingh (3.08%) and Rajshahi (3.14%) (Table 1).

Marital status showed a strong association: women who were separated, divorced, or widowed had markedly higher depressive symptoms (11.39%) compared to currently married women (4.52%) (p < 0.001). Similarly, women whose husbands had less than secondary education had higher prevalence (5.63% vs. 4.17%, p < 0.001).

Autonomy and access-related factors were also important. Women without decision-making autonomy had higher depressive symptoms (5.63% vs. 4.65%, p = 0.028). Those reporting distance as a barrier to healthcare (5.66% vs. 4.13%) and those facing healthcare access problems (5.36% vs. 4.00%) showed significantly higher prevalence (p < 0.001 for both).

Household and environmental conditions revealed notable disparities. Women living in households where cooking occurred indoors (6.30%) or outdoors (6.65%) had higher depressive symptoms compared to those using a separate building (4.25%) (p < 0.001). Women without livestock (5.64% vs. 4.30%, p < 0.001) and those without agricultural land (6.71% vs. 4.62%, p < 0.001) also had higher prevalence.

Reproductive factors were significantly associated with depressive symptoms. Women who had ever experienced pregnancy termination had higher prevalence (5.84% vs. 4.50%, p = 0.001). Likewise, women without children under five years had higher depressive symptoms (5.45% vs. 3.91%, p < 0.001).

Among continuous variables, women with depressive symptoms were older (32.50 vs. 30.72 years, p < 0.001), had higher wife-beating attitude scores (1.06 vs. 0.39, p < 0.001), and had slightly more living children (1.97 vs. 1.87, p = 0.016). No significant differences were observed for education, household head age, residence, employment status, media exposure, or household crowding.

### Feature Engineering and Selection

The feature selection process identified a reduced subset of predictors associated with depressive symptoms. Elastic Net retained 29 variables with non-zero coefficients at the optimal λ, while Random Forest and XGBoost each identified the top 15 predictors based on importance metrics (Fig. 4). To enhance robustness and reduce model-specific bias, the final feature set was defined as the intersection of predictors consistently identified across all three methods, resulting in 16 variables included in the final model [39].

**Fig. 4.**
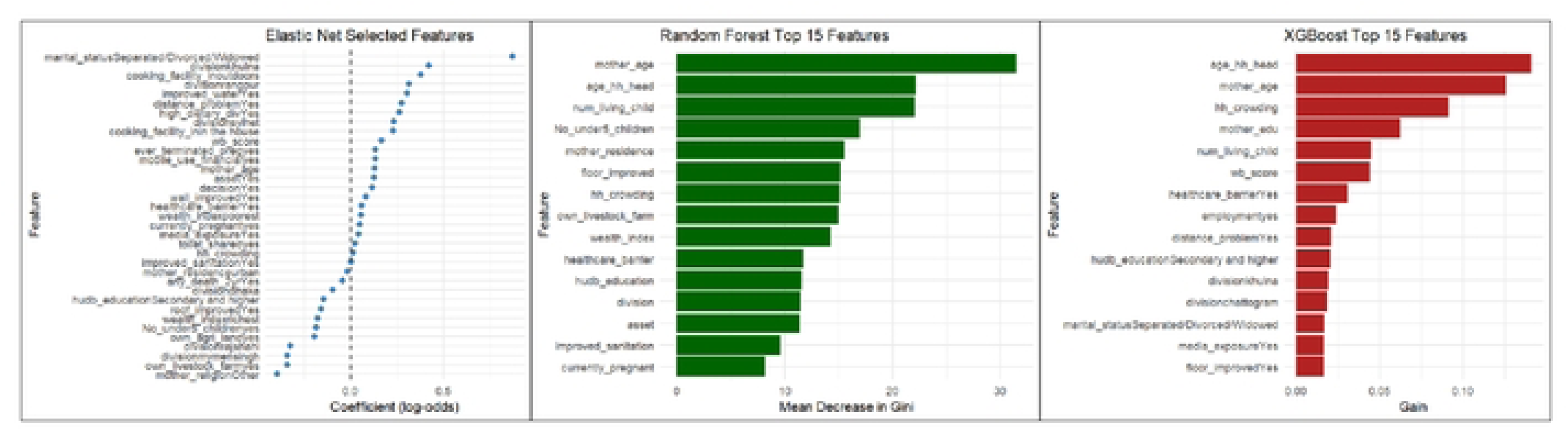
Feature importance and selection across models. (a) Elastic Net–selected predictors with non-zero coefficients, shown on the log-odds scale. (b) Top 15 predictors ranked by mean decrease in Gini from the Random Forest model, capturing nonlinear effects and interactions. (c) Top 15 predictors ranked by gain from the XGBoost model, reflecting their relative contribution to loss reduction.

Key predictors such as age, marital status, healthcare barriers, and geographic division were consistently identified across both traditional statistical analyses and machine learning approaches, highlighting their robustness. Additionally, machine learning models emphasized the importance of household and environmental factors, suggesting potential nonlinear relationships and interactions.

### Model Performance

Table 2 presents the predictive performance of the proposed bagging ensemble approach using different weak learners and their optimal voting thresholds. Across all models, the normalized Matthews correlation coefficient (normMCC) ranged from 0.543 to 0.557, indicating moderate and relatively stable discrimination under severe class imbalance.

**Table 2.** Predictive performance of the proposed ensemble method with different weak learners for predicting depressive symptoms (Th)

| Model | Best threshold | Accuracy | Sensitivity | Specificity | F1-score | normMCC |
| --- | --- | --- | --- | --- | --- | --- |
| EN | 16 | 0.701 | 0.534 | 0.710 | 0.147 | 0.557 |
| RF | 15 | 0.724 | 0.494 | 0.735 | 0.147 | 0.555 |
| SVM | 19 | 0.803 | 0.378 | 0.824 | 0.156 | 0.556 |
| KNN | 17 | 0.800 | 0.333 | 0.824 | 0.138 | 0.543 |
| GBM | 14 | 0.693 | 0.526 | 0.701 | 0.141 | 0.553 |

EN = Elastic Net Logistic Regression; RF = Random Forest; SVM = Support Vector Machine; KNN = K-nearest neighbors; GBM = Gradient Boosting Machine, normMCC = normalized Matthews correlation coefficient.

The EN ensemble achieved the highest normMCC (0.557) and the highest sensitivity (0.534), suggesting comparatively better identification of women with depressive symptoms, although its specificity (0.710) was lower than that of SVM and KNN. The Random Forest (RF) ensemble showed the most balanced performance overall, with accuracy of 0.724, sensitivity of 0.494, specificity of 0.735, and normMCC of 0.555.

The SVM ensemble attained the highest accuracy (0.803) and specificity (0.824), but exhibited lower sensitivity (0.378), indicating a stronger tendency to correctly classify non-depressed individuals at the expense of missing a larger proportion of depressed cases. A similar pattern was observed for the KNN ensemble, which achieved high accuracy (0.800) and specificity (0.824) but the lowest sensitivity (0.333) and normMCC (0.543).

The GBM ensemble demonstrated a more balanced trade-off between sensitivity (0.526) and specificity (0.701), yielding a normMCC of 0.553. Overall, the ensemble framework produced consistent performance across different base learners, with EN, RF, and GBM providing more favorable sensitivity–specificity trade-offs for depressive symptoms prediction than SVM and KNN.

We compared the predictive performance of the proposed bagging ensemble models with oversampling-based models across five algorithms (Table 3). In general, ensemble learning improved F1-score and normMCC for most classifiers, indicating more balanced performance under class imbalance.

**Table 3.** Comparison of predictive performance metrics for oversampling-based and proposed ensemble models across five algorithms.

| Model | Oversample |  |  |  |  | Proposed Ensemble model |  |  |  |  |
| --- | --- | --- | --- | --- | --- | --- | --- | --- | --- | --- |
|  | Accuracy | Sensitivity | Specificity | F1-score | norm MCC | Accuracy | Sensitivity | Specificity | F1-score | norm MCC |
| EN | 0.625 | 0.598 | 0.626 | 0.133 | 0.549 | 0.701 | 0.534 | 0.710 | 0.147 | 0.557 |
| RF | 0.835 | 0.237 | 0.865 | 0.121 | 0.531 | 0.724 | 0.494 | 0.735 | 0.147 | 0.555 |
| SVM | 0.723 | 0.426 | 0.738 | 0.129 | 0.539 | 0.803 | 0.378 | 0.824 | 0.156 | 0.556 |
| KNN | 0.721 | 0.410 | 0.737 | 0.124 | 0.535 | 0.800 | 0.333 | 0.824 | 0.138 | 0.543 |
| GBM | 0.877 | 0.217 | 0.910 | 0.145 | 0.546 | 0.693 | 0.526 | 0.701 | 0.141 | 0.553 |

EN = Elastic Net Logistic Regression; RF = Random Forest; SVM = Support Vector Machine; KNN = K-nearest neighbors; GBM = Gradient Boosting Machine. normMCC = normalized Matthews correlation coefficient.

For the EN model, the ensemble approach improved accuracy (0.701 vs. 0.625), specificity (0.710 vs. 0.626), F1-score (0.147 vs. 0.133), and normMCC (0.557 vs. 0.549), despite a modest reduction in sensitivity (0.534 vs. 0.598). These results suggest that ensemble learning enhanced overall discrimination while slightly reducing the detection rate of depressed cases.

The RF ensemble showed substantial gains over its oversampling counterpart, particularly in sensitivity (0.494 vs. 0.237), F1-score (0.147 vs. 0.121), and normMCC (0.555 vs. 0.531), although accuracy and specificity decreased. This indicates that the ensemble strategy markedly improved minority-class detection for tree-based models.

For SVM, the ensemble approach increased accuracy (0.803 vs. 0.723), specificity (0.824 vs. 0.738), F1-score (0.156 vs. 0.129), and normMCC (0.556 vs. 0.539), while sensitivity declined slightly (0.378 vs. 0.426). A similar pattern was observed for KNN, where ensemble learning improved accuracy, specificity, F1-score, and normMCC but reduced sensitivity relative to oversampling.

In contrast, the GBM ensemble substantially increased sensitivity (0.526 vs. 0.217) and normMCC (0.553 vs. 0.546) compared with oversampling, although accuracy and specificity were lower. This suggests that ensemble learning particularly benefited GBM in detecting depressed cases.

Taken together, these findings demonstrate that bagging ensemble models generally outperformed oversampling-based models in terms of balanced classification performance, especially for RF and GBM algorithms. While oversampling tended to increase sensitivity for some models, ensemble learning provided a more favorable trade-off between sensitivity and specificity, resulting in higher F1-scores and normMCC values for most classifiers.

### Threshold Optimization for Bagging Ensemble Models

Fig. 5 illustrates how classification performance metrics vary with the decision threshold (Th) for each ensemble model (EN, GBM, SVM, KNN, and RF). Across all models, increasing the threshold leads to a consistent trade-off between sensitivity and specificity. Specifically, sensitivity decreases monotonically as Th increases, while specificity and overall accuracy increase. This pattern reflects stricter voting requirements for classifying an observation as depressed, thereby reducing false positives at the expense of missing true cases.

**Fig. 5.**
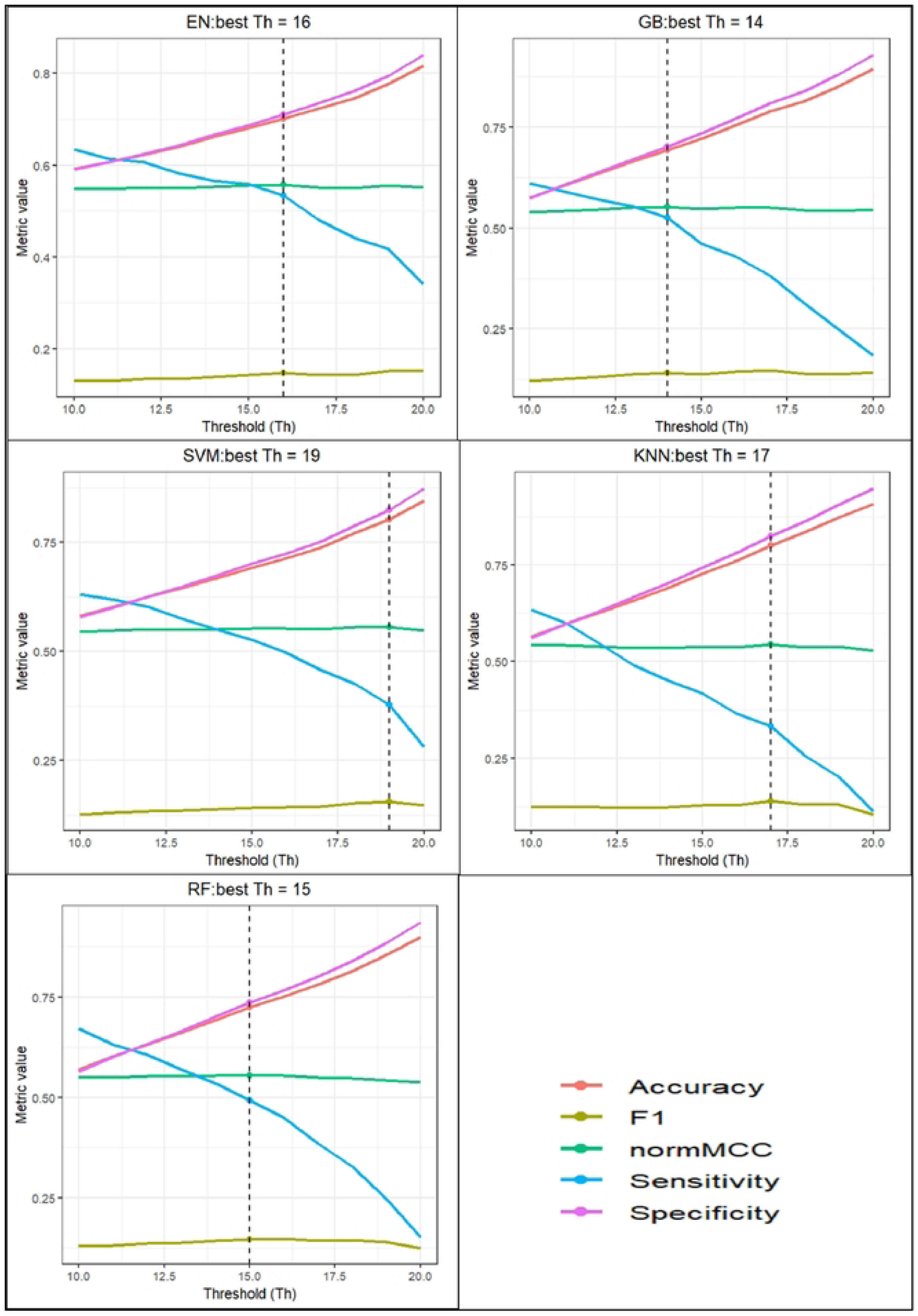
Performance of bagging ensemble models across varying decision thresholds (Th). For each classifier (Elastic Net (EN), Gradient Boosting (GB), Support Vector Machine (SVM), K-nearest neighbors (KNN), and RF: Random Forest), accuracy, sensitivity, specificity, F1-score, and normalized Matthews correlation coefficient (normMCC) are plotted against the threshold value

The normalized Matthews correlation coefficient (normMCC) exhibits a unimodal pattern for most models, peaking at intermediate threshold values. These peaks correspond to the model-specific optimal thresholds (indicated by vertical dashed lines), which balance sensitivity and specificity under class imbalance. For EN and GBM, the optimal thresholds (Th = 16 and Th = 14, respectively) coincide with relatively high sensitivity and moderate specificity, suggesting improved minority-class detection. In contrast, SVM and KNN achieve their optimal performance at higher thresholds (Th = 19 and Th = 17), reflecting a stronger emphasis on specificity and overall accuracy. The RF model attains its optimal threshold at Th = 15, yielding a balanced trade-off between sensitivity and specificity.

Across all models, the F1-score changes more gradually with threshold than sensitivity or specificity, indicating limited improvement in precision–recall balance beyond the selected optimal thresholds. Overall, these results highlight the importance of threshold tuning in bagging ensemble models and demonstrate that the optimal decision threshold varies by learning algorithm, reflecting differences in model structure and error trade-offs.

## Discussion

In this study, we developed and evaluated the performance of bagging-based ensemble machine learning models and compared with oversampling-based classical models for predicting depressive symptoms among reproductive-aged women in Bangladesh using highly imbalanced BDHS 2022 data. Our findings highlight both the methodological challenges of modeling rare mental health outcomes in population-based surveys and the advantages of ensemble learning under severe class imbalance.

Across all five algorithms, the proposed bagging ensemble framework generally outperformed oversampling-based models in terms of normMCC, metric that provide more reliable assessments of predictive performance in imbalanced classification settings than accuracy alone [40]. While oversampling approaches tended to improve sensitivity for certain classifiers, this improvement was often achieved at the cost of reduced specificity and overall discrimination. In contrast, ensemble learning produced a more favorable balance between detecting depressed cases and limiting false-positive classifications, resulting in more stable and interpretable performance (Fig. 6).

From a methodological perspective, this study contributes to the growing literature on applying machine learning to population-based survey data by demonstrating that extreme class imbalance in DHS-type datasets poses unique challenges that are not fully addressed by conventional resampling approaches. The proposed bagging framework, combined with threshold optimization, provides a flexible strategy to improve minority-class detection while preserving the original data structure. In addition, the use of normMCC as a primary evaluation metric ensures a more balanced and robust assessment of model performance under severe imbalance, which is particularly important for rare mental health outcomes.

Among the evaluated classifiers, the Random Forest ensemble demonstrated the most balanced overall performance. Compared with its oversampling-based counterpart, the ensemble Random Forest substantially improved sensitivity, F1-score, and normMCC while maintaining relatively high specificity. This improvement likely reflects the ability of bagging to reduce variance and mitigate class dominance when training tree-based models on balanced subsets of the data [41]. Random Forest models are well suited to this strategy because they naturally capture nonlinear effects and interactions among predictors while benefiting from diversity across bootstrap samples[42].

The Gradient Boosting ensemble also showed marked improvement in minority-class detection, with sensitivity more than doubling relative to the oversampling-based Gradient Boosting model. This gain was accompanied by an increase in normMCC, indicating improved correlation between predicted and observed outcomes under imbalance. These findings are consistent with prior work showing that ensemble resampling strategies can enhance the robustness of boosting methods when outcomes are rare [16].

For Elastic Net logistic regression, ensemble learning improved accuracy, specificity, F1-score, and normMCC despite a modest reduction in sensitivity. This suggests that combining regularized linear models with a bagging strategy can improve overall discrimination by reducing the dominance of the majority class while maintaining interpretability. The Support Vector Machine ensemble achieved the highest accuracy and specificity but lower sensitivity, indicating a stronger tendency to classify women as non-depressed. This pattern reflects the known behavior of margin-based classifiers under imbalance, where decision boundaries tend to favor the majority class unless explicitly penalized [15, 43].

K-Nearest Neighbors showed limited benefit from ensemble learning in terms of sensitivity, although accuracy and specificity increased. Because KNN relies on local neighborhood structure, it is highly sensitive to class distribution and density, which may limit its responsiveness to resampling strategies compared with tree-based or regularized models [44].

Threshold optimization further demonstrated the flexibility and robustness of the ensemble framework. Across all classifiers, increasing the voting threshold produced the expected trade-off between sensitivity and specificity, with higher thresholds favoring specificity and lower thresholds favoring sensitivity. The normMCC exhibited a unimodal pattern, peaking at intermediate threshold values that balanced these competing objectives. Importantly, normMCC remained relatively stable across a wide range of thresholds, suggesting that ensemble performance was not overly sensitive to the choice of voting rule. This property is desirable for public health applications, where different operational thresholds may be required depending on whether the priority is early case detection or minimizing false positives.

Despite these methodological advantages, the absolute predictive performance of all models remained modest, with normMCC values ranging from approximately 0.53 to 0.56. This likely reflects the inherent difficulty of predicting depressive symptoms using cross-sectional survey data, where mental health outcomes are influenced by unmeasured psychosocial, environmental, and biological factors. In addition, the low prevalence of depression in the study population imposes statistical limits on achievable performance. Similar moderate discrimination has been reported in other population-based prediction studies using machine learning methods [45].

From a public health perspective, these findings suggest that bagging-based ensemble learning offers a more reliable and transparent framework than synthetic oversampling for depression risk prediction in highly imbalanced epidemiological data. Rather than maximizing sensitivity alone, the ensemble approach yields balanced performance that may be better suited for large-scale screening, surveillance, and prioritization of high-risk groups.

## Limitations

Several limitations should be acknowledged. First, depressive symptoms were measured using a screening instrument rather than a clinical diagnosis, and misclassification is possible due to reporting bias or transient symptoms at the time of interview. Second, the BDHS does not capture important psychosocial and contextual factors such as social support, intimate partner violence, life events, or prior mental health history, which are known to influence depression risk and may constrain achievable predictive performance.

Third, although the BDHS employs a complex survey design, sampling weights, stratification, and clustering were not incorporated into model development. As a result, predicted probabilities may not fully reflect nationally representative population estimates. However, this study focused on individual-level risk prediction rather than population-level inference, where the use of sampling weights is more critical. In predictive modeling, performance is evaluated based on out-of-sample accuracy rather than unbiased estimation of population parameters, and prior research suggests that omission of weights often has limited impact on classification performance.

Fourth, although ensemble learning improved balanced performance, sensitivity remained modest, indicating that a substantial proportion of cases would still be missed if the models were used for screening purposes. Finally, the feature selection strategy prioritized stability across models, which may have excluded potentially relevant predictors and limited model flexibility. Future studies should explore the integration of longitudinal data, richer psychosocial measures, and survey-weighted or cost-sensitive learning approaches to enhance both predictive accuracy and population-level applicability.

## Conclusion

Bagging-based ensemble learning provided more balanced and robust predictive performance than oversampling-based methods for identifying depressive symptoms in highly imbalanced population survey data. Although overall discrimination was moderate, the ensemble framework improved normMCC and achieved a more favorable trade-off between sensitivity and specificity across multiple classifiers. These findings support the use of aggregation-based ensemble strategies for depression risk stratification in epidemiological studies and suggest potential utility for targeted screening and population-level mental health surveillance in resource-limited settings.

## Data Availability

The data set used and analyzed in this study is freely available at the Demographic and Health Surveys (DHS) program website https://dhsprogram.com/data/available-datasets.cfm.

---

Supplementary File 1: Variables details

Supplementary File 2: Hyperparameters optimization

Supplementary File 3: TRIPODAI_checklist

## Reference

1. Zhao L, Lou Y, Tao Y, Wang H, Xu N. Global, regional and national burden of depressive disorders in adolescents and young adults, 1990–2021: systematic analysis of the global burden of disease study 2021. Frontiers in Public Health. 2025;13:1599602.

2. Dai F, Cai Y, Chen M, Dai Y. Global trends of depressive disorders among women of reproductive age from 1990 to 2021: a systematic analysis of burden, sociodemographic disparities, and health workforce correlations. BMC Psychiatry. 2025;25(1):263. Epub 20250320. doi: 10.1186/s12888-025-06697-4. PubMed PMID: 40114132; PubMed Central PMCID: PMCPMC11924784.

3. Gelaye B, Rondon MB, Araya R, Williams MA. Epidemiology of maternal depression, risk factors, and child outcomes in low-income and middle-income countries. Lancet Psychiatry. 2016;3(10):973–82. Epub 20160917. doi: 10.1016/s2215-0366(16)30284-x. PubMed PMID: 27650773; PubMed Central PMCID: PMCPMC5155709.

4. Arafat SMY, Rajkumar RP. Mental disorders during pregnancy and postpartum in Bangladesh: A narrative review. Health Sci Rep. 2024;7(9):e70027. Epub 20240828. doi: 10.1002/hsr2.70027. PubMed PMID: 39210993; PubMed Central PMCID: PMCPMC11358212.

5. Hossain AT, Rahman MH, Manna RM, Akter E, Islam SH, Hossain MA, et al. Enhancing Access to Mental Health Services for Antepartum and Postpartum Women Through Telemental Health Services at Wellbeing Centers in Selected Health Facilities in Bangladesh: Implementation Research. JMIR Pediatr Parent. 2025;8:e65912. Epub 3.1.2025. doi: 10.2196/65912.

6. Moitra M, Santomauro D, Collins PY, Vos T, Whiteford H, Saxena S, et al. The global gap in treatment coverage for major depressive disorder in 84 countries from 2000-2019: A systematic review and Bayesian meta-regression analysis. PLoS Med. 2022;19(2):e1003901. Epub 20220215. doi: 10.1371/journal.pmed.1003901. PubMed PMID: 35167593; PubMed Central PMCID: PMCPMC8846511.

7. National Institute of Population Research and Training (NIPORT) and ICF. Bangladesh Demographic and Health Survey 2022: Final Report. Dhaka, Bangladesh, and Rockville, Maryland, USA: NIPORT and ICF: 2024.

8. Yu C, Kong X, Yu W, Ni X, Chen J, Liao X. Machine learning models for predicting the risk of depressive symptoms in Chinese college students. Front Psychiatry. 2025;16:1648585. Epub 20250805. doi: 10.3389/fpsyt.2025.1648585. PubMed PMID: 40838255; PubMed Central PMCID: PMCPMC12361154.

9. Xin B, Guo B, Li Q, Li M, Tong F, Wu Y, et al. Machine learning-based risk prediction models for depressive symptoms in Chinese community-dwelling individuals with multimorbidity: A longitudinal study. Journal of Affective Disorders. 2025:120075.

10. Andersson S, Bathula DR, Iliadis SI, Walter M, Skalkidou A. Predicting women with depressive symptoms postpartum with machine learning methods. Scientific reports. 2021;11(1):7877.

11. Arias de la Torre J, Vilagut G, Ronaldson A, Bakolis I, Dregan A, Navarro-Mateu F, et al. Reconsidering the Use of Population Health Surveys for Monitoring of Mental Health. JMIR Public Health Surveill. 2023;9:e48138. Epub 23.11.2023. doi: 10.2196/48138. PubMed PMID: 37995112.

12. Miah MS, Ullah MO. Identifying the Determinants of Depression among Ever Married Women in Bangladesh: A Machine Learning Approach. International Journal of Statistical Sciences. 2025;25(2):33–46.

13. Sabouri Z, Moustati I, Gherabi N, Amnai M. Interpretable Machine Learning Framework for Early Depression Detection Using Socio-Demographic Features with Dual Feature Selection and SMOTE. Informatica. 2025;49(4).

14. Chawla NV, Bowyer KW, Hall LO, Kegelmeyer WP. SMOTE: synthetic minority over-sampling technique. Journal of artificial intelligence research. 2002;16:321–57.

15. He H, Garcia E. Learning from Imbalanced Data IEEE Transactions on Knowledge and Data Engineering. vol; 2009.

16. Wang J, Black M, Rankin D, Wallace J, Hughes C, Hoey L, et al. A Bagging Ensemble Machine Learning Method for Imbalanced Data to Predict Anxiety Disorders and Analysis of Risk Factors in Older People: Observational Study. Artificial Intelligence in Health. 2025:1–22.

17. Breiman L. Bagging Predictors. Machine Learning. 1996;24(2):123–40. doi: 10.1023/A:1018054314350.

18. Chubak J, Pocobelli G, Weiss NS. Tradeoffs between accuracy measures for electronic health care data algorithms. J Clin Epidemiol. 2012;65(3):343–9.e2. Epub 20111223. doi: 10.1016/j.jclinepi.2011.09.002. PubMed PMID: 22197520; PubMed Central PMCID: PMCPMC3264740.

19. Wahid SS, Raza WA, Mahmud I, Kohrt BA. Climate-related shocks and other stressors associated with depression and anxiety in Bangladesh: a nationally representative panel study. The lancet planetary health. 2023;7(2):e137–e46.

20. Chowdhury AN, Sayanti Ghosh SG, Debasish Sanyal DS. Bengali adaptation of brief patient health questionnaire for screening depression at primary care. 2004.

21. Thomas T, Rajabi E. A systematic review of machine learning-based missing value imputation techniques. Data Technologies and Applications. 2021;55(4):558–85. doi: 10.1108/dta-12-2020-0298.

22. Bermingham ML, Pong-Wong R, Spiliopoulou A, Hayward C, Rudan I, Campbell H, et al. Application of high-dimensional feature selection: evaluation for genomic prediction in man. Scientific reports. 2015;5(1):10312.

23. Jović A, Brkić K, Bogunović N, editors. A review of feature selection methods with applications. 2015 38th international convention on information and communication technology, electronics and microelectronics (MIPRO); 2015: Ieee.

24. Liu W, Li Q. An efficient elastic net with regression coefficients method for variable selection of spectrum data. PloS one. 2017;12(2):e0171122.

25. Lei S, editor A feature selection method based on information gain and genetic algorithm. 2012 international conference on computer science and electronics engineering; 2012: IEEE.

26. Chen C, Zhang Q, Yu B, Yu Z, Lawrence PJ, Ma Q, et al. Improving protein-protein interactions prediction accuracy using XGBoost feature selection and stacked ensemble classifier. Computers in biology and medicine. 2020;123:103899.

27. Hong S, Hong S, Oh E, Lee WJ, Jeong WK, Kim K. Development of a flexible feature selection framework in radiomics-based prediction modeling: Assessment with four real-world datasets. Scientific Reports. 2024;14(1):29297. doi: 10.1038/s41598-024-80863-8.

28. Kim M-h, Banerjee S, Park SM, Pathak J, editors. Improving risk prediction for depression via elastic net regression-results from korea national health insurance services data. AMIA annual symposium proceedings; 2017.

29. Rigatti SJ. Random forest. Journal of insurance medicine. 2017;47(1):31–9.

30. Pisner DA, Schnyer DM. Support vector machine. Machine learning: Elsevier; 2020. p. 101–21.

31. Hodge VJ, Austin J. A binary neural k-nearest neighbour technique. Knowledge and Information Systems. 2005;8(3):276–91.

32. Bahad P, Saxena P, editors. Study of adaboost and gradient boosting algorithms for predictive analytics. International Conference on Intelligent Computing and Smart Communication 2019: Proceedings of ICSC 2019; 2019: Springer.

33. Salam MA, Islam MM, Karim MR. Machine learning algorithms for predicting and identifying the influencing predictors of antenatal care visits among women in Bangladesh: Evidence from BDHS 2022 data. Plos one. 2025;20(10):e0324226.

34. Chicco D, Tötsch N, Jurman G. The Matthews correlation coefficient (MCC) is more reliable than balanced accuracy, bookmaker informedness, and markedness in two-class confusion matrix evaluation. BioData mining. 2021;14(1):13.

35. MacNell N, Feinstein L, Wilkerson J, Salo PM, Molsberry SA, Fessler MB, et al. Implementing machine learning methods with complex survey data: Lessons learned on the impacts of accounting sampling weights in gradient boosting. PLoS One. 2023;18(1):e0280387. Epub 20230113. doi: 10.1371/journal.pone.0280387. PubMed PMID: 36638125; PubMed Central PMCID: PMCPMC9838837.

36. Kaombe TM, Hamuza GA. Impact of ignoring sampling design in the prediction of binary health outcomes through logistic regression: evidence from Malawi demographic and health survey under-five mortality data; 2000-2016. BMC Public Health. 2023;23(1):1674. Epub 20230831. doi: 10.1186/s12889-023-16544-4. PubMed PMID: 37653375; PubMed Central PMCID: PMCPMC10469829.

37. Naznin S, Uddin MJ, Kabir A. Identifying determinants of under-5 mortality in Bangladesh: A machine learning approach with BDHS 2022 data. Plos one. 2025;20(6):e0324825.

38. Collins GS, Moons KG, Dhiman P, Riley RD, Beam AL, Van Calster B, et al. TRIPOD+ AI statement: updated guidance for reporting clinical prediction models that use regression or machine learning methods. bmj. 2024;385.

39. Levandowski BA, Pro GC, Rietberg-Miller SB, Camplain R. We are complex beings: comparison of statistical methods to capture and account for intersectionality. BMJ Open. 2024;14(1):e077194. Epub 20240130. doi: 10.1136/bmjopen-2023-077194. PubMed PMID: 38296287; PubMed Central PMCID: PMCPMC10828873.

40. Chicco D, Jurman G. The advantages of the Matthews correlation coefficient (MCC) over F1 score and accuracy in binary classification evaluation. BMC Genomics. 2020;21(1):6. Epub 20200102. doi: 10.1186/s12864-019-6413-7. PubMed PMID: 31898477; PubMed Central PMCID: PMCPMC6941312.

41. Galar M, Fernandez A, Barrenechea E, Bustince H, Herrera F. A review on ensembles for the class imbalance problem: bagging-, boosting-, and hybrid-based approaches. IEEE Transactions on Systems, Man, and Cybernetics, Part C (Applications and Reviews). 2011;42(4):463–84.

42. Alvarez Espezua CB, Cruz de la Cruz JE, Apaza Davila FA, Cruz de la Cruz TD, Huaquipaco Encinas S, Mamani Machaca WA, editors. Classification of Depression and Anxiety with Machine Learning Applying Random Forest Models. Proceedings of the 2024 5th International Conference on Intelligent Medicine and Health; 2024.

43. Feng W, Huang W, Ren J. Class imbalance ensemble learning based on the margin theory. Applied Sciences. 2018;8(5):815.

44. Gøttcke JMN, Zimek A, editors. Handling class imbalance in k-nearest neighbor classification by balancing prior probabilities. International Conference on Similarity Search and Applications; 2021: Springer.

45. Zheng Y, Zhang T, Yang S, Wang F, Zhang L, Liu Y. Using machine learning to predict the probability of incident 2-year depression in older adults with chronic diseases: a retrospective cohort study. BMC Psychiatry. 2024;24(1):870. Epub 20241202. doi: 10.1186/s12888-024-06299-6. PubMed PMID: 39623372; PubMed Central PMCID: PMCPMC11610371.

